# Estimating the potential malaria morbidity and mortality avertable by the President’s Malaria Initiative in 2025: a geospatial modelling analysis

**DOI:** 10.1101/2025.02.28.25323072

**Authors:** Tasmin L. Symons, Jailos Lubinda, Michael McPhail, Adam Saddler, Mauricio van den Berg, Hunter Baggen, Yuval Berman, Sarah Hafsia, Rubi Jayaseelen, Punam Amratia, Annie Browne, Ewan Cameron, Camilo Vargas-Ruiz, Susan F. Rumisha, Nick Golding, Daniel J. Weiss, Peter W. Gething

**Affiliations:** School of Population Health, Curtin University, Bentley, Western Australia; Malaria Atlas Project, The Kids Research Institute Australia, Nedlands, Western Australia; Malaria Atlas Project, Ifakara Health Institute, Dar es Salaam, Tanzania; School of Physics, Mathematics and Computing, University of Western Australia, Western Australia; Melbourne School of Population and Global Health, University of Melbourne, Victoria

## Abstract

**Background:** Since its inception in 2005, the President’s Malaria Initiative (PMI) has played a major role in the reductions in malaria morbidity and mortality witnessed across Africa. With the status of PMI funding and operations currently uncertain, this study aimed to quantify the impact that a fully-functioning PMI would have on malaria cases and deaths in Africa during 2025.

**Methods:** We combined detailed spatio-temporal information on planned 2025 PMI and non-PMI malaria commodity procurement and distribution in Africa with spatio-temporal Bayesian models of intervention coverage and *Plasmodium falciparum* transmission and burden in Africa. By comparing coverage scenarios with and without planned PMI contributions we estimated the number of malaria cases and deaths PMI would avert in 2025.

**Findings:** We estimated that business-as-usual PMI contributions to vector control, seasonal chemoprevention, and routine malaria treatment in Africa would avert in 2025 13·6M (95% uncertainty interval 11·4M – 16·4M) malaria cases and 104,000 (69,000 – 161,000) deaths. This represents 11·3% (9·9 – 12·9%) and 37·5% (34·2 – 41·5%), respectively, of the total burden of malaria morbidity and mortality in PMI’s focus geographies across 27 African countries. These estimates do not account for the additional impact of PMI-supported provision of diagnostics or severe case management commodities, nor preventive treatment for pregnant women which would add further to the averted burden.

**Interpretation:** PMI investment in supporting procurement and distribution of malaria control commodities would translate directly into millions of malaria cases averted and a hundred thousand lives saved across its focus geographies in Africa across 2025.

**Research in context:** *Evidence before this study:* We searched PubMed on 25 February 2025 with the search term “President’s Malaria Initiative” AND (impact OR evaluation) AND (burden OR malaria cases OR malaria deaths). 102 results were returned, of which four directly addressed the question of PMI-attributable impact on malaria burden across all its focus geographies. One focussed solely on case-management and did not provide quantitative results^1^. A study focussing on mortality found a strong association between PMI investment and declines in all-cause infant mortality^2^. Another study used a generalised estimating equation approach to assess PMI’s impact, concluding a strong association between PMI spending and reductions in malaria burden^3^. A 2017 study used a mechanistic malaria model to estimate PMI’s historic contribution to reductions in malaria morbidity and mortality, with future projections to 2020^4^.

*Added value of this study:* In this study we synthesised the most up-to-date information of all-funder volumes of key malaria control interventions (ITN, IRS, ACT, SMC) with PMI data on planned volumes and spatial targeting of funding in 2025 to derive near-real-time projections of malaria control intervention coverage in Africa under two scenarios: a business-as-usual scenario in which PMI commodities procured and distributed as previously planned versus a ‘no-PMI’ scenario in which PMI funding and technical assistance is absent. These scenario-based estimates of coverage were propagated through an empirical geospatial malaria model, coupled to mechanistic models of clinical incidence, in order to estimate the number of malaria infections, clinical cases, and deaths PMI activities could expect to avert in Africa in 2025.

*Implications of all available evidence:* PMI is a key contributor to malaria control efforts in Africa, and as such its impact on averting childhood mortality has been demonstrated repeatedly. Here, using empirically-derived estimates of malaria control impact, combined with detailed scenarios of intervention coverage with and without PMI support, we isolated the contemporary impact of PMI activities on malaria morbidity and mortality in Africa in 2025. Our findings suggest that PMI support is critical to maintaining suppression of malaria transmission in its focus geographies.

## Introduction

Over the past 20 years, malaria control has greatly reduced malaria transmission, cases, and deaths in Africa^5,6^. A catalyst of this achievement has been the growth in international development assistance for health, including the launch in 2002 of the Global Fund followed in 2005 by the US President’s Malaria Initiative (PMI). By 2023 PMI supported malaria control in 31 countries (27 in Africa), and disbursed USD$777·7M^7^. That investment enabled, in the 2023 US financial year, the procurement of 36·8M insecticide treated bednets (ITNs), 63·3M doses of the effective antimalarial drug artemisinin-based combination therapy (ACT), indoor residual spraying (IRS) of insecticides to protect 15·5M people, and 48M treatments of seasonal malaria chemoprevention (SMC) to protect children in locations with intense malaria seasonality_7._ Most of this financial support was directed to the highest burden regions of malaria-endemic Africa^6,7^. In addition to core vector control and treatment, PMI has also supported the large-scale deployment of preventive treatment for pregnant women, the scale-up of rapid diagnostic tests (RDTs) to support accurate point-of-care diagnosis, and other wide-ranging technical support to health personnel working in malaria-endemic countries. The immediate and longer-term future of PMI is currently uncertain, and the extent to which it’s planned support for malaria control across Africa in 2025 will eventuate is not clear. Given the large proportional contribution of PMI to overall malaria control efforts in the highest-burden regions, the presence or absence of PMI financing and operations has the potential to significantly impact total malaria burden in Africa. Estimates of the overall potential impact of PMI in 2025, how this varies by country, and the relative contributions of different intervention tools, can therefore help inform ongoing decision making at international and national levels.

Various approaches have been developed to estimate - retrospectively or prospectively - the malaria burden averted by investments in malaria control efforts^8,9^. An initial challenge is to understand how those investments have translated into coverage of malaria control interventions. A common method is to assume that funds invested translate proportionally into coverage achieved, enabling coverage to be predicted using simple arithmetic based on commodity unit costs multiplied by funds available. While reasonable for some interventions, this assumption is not always valid and evidence demonstrates, for example, that incremental increases in coverage of ITNs becomes progressively more expensive to achieve above a certain threshold^10,11^. In this study, we deploy a range of coverage models that more explicitly link investments to commodity volumes, to deployment, and ultimately to population coverage. The second challenge is to estimate the likely impact interventions have on malaria at a given level of coverage. In this study we build on the modelling architecture of the Malaria Atlas Project (MAP)^12^ used to produce annually updated estimates of malaria burden and trends for the WHO World Malaria Report^6^ and Global Burden of Disease study^13^. We use this framework to generate modelled estimates of the potential of PMI to avert malaria cases and deaths in 2025. We define a business-as-usual scenario in which we reconstruct malaria control commodity procurement and distribution as previously planned for 2025 and compare this to a scenario in which activities supported by PMI do not take place.

## Methods

Figure 1 provides a schematic overview of our analysis, which comprised the following steps. First, we assembled data on past and planned malaria control commodity procurement and distribution from both PMI and non-PMI funding sources and focusing on ITNs, ACTs, SMC, and IRS. Second, we used coverage models to estimate subnational coverage of each of these interventions in 2025 in scenarios with and without contributions from PMI. Third, we used an established Bayesian geostatistical model to predict infection prevalence across Africa in 2025 under both scenarios and, fourth, converted these into corresponding estimates of malaria clinical incidence and mortality. These steps are described in more detail below and with supporting methodological descriptions in the appendix.

**Figure 1.**
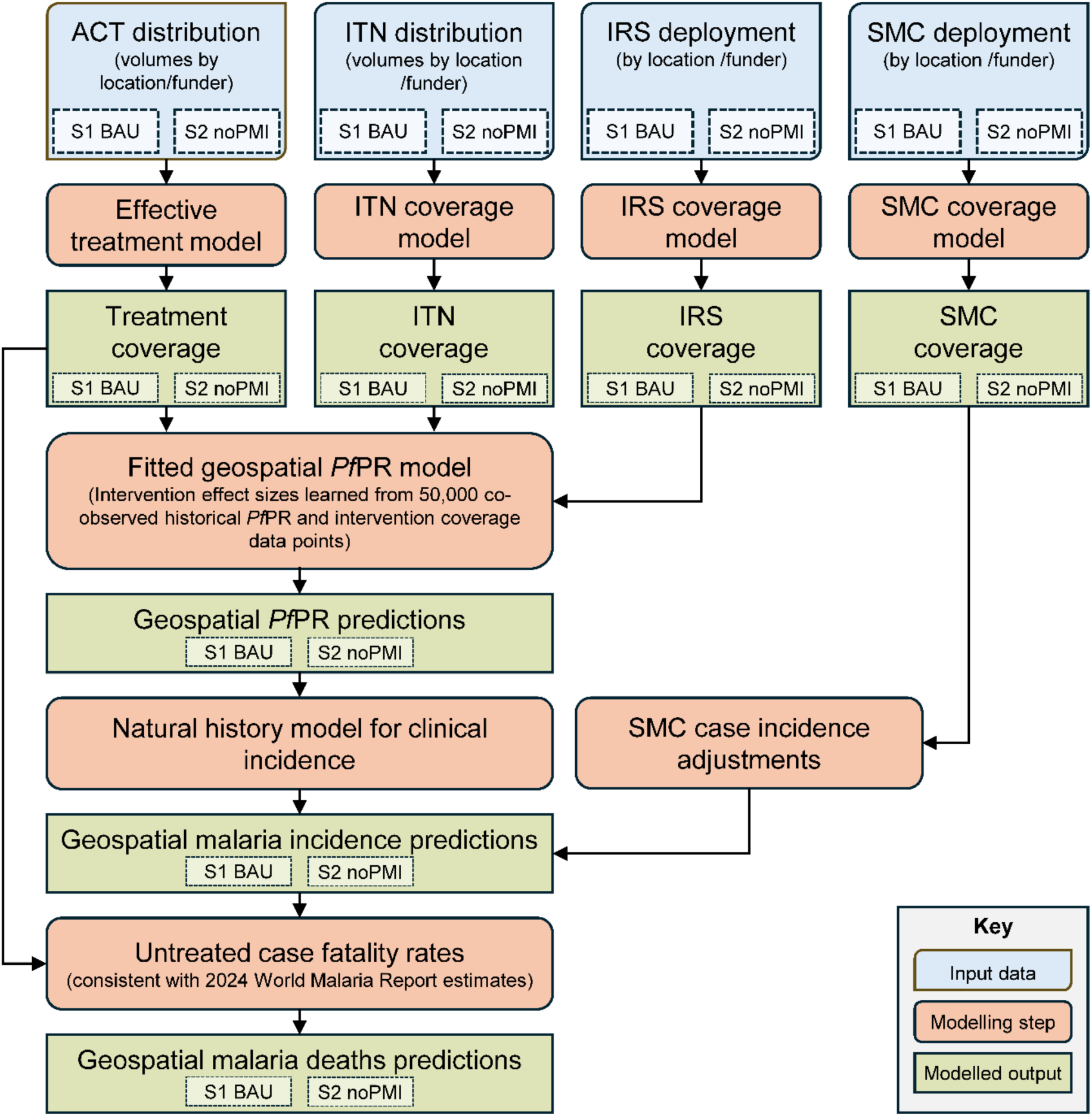
Schematic summary of methodological framework. Blue rectangles represent input datasets, some of which are modelled results from preceding analyses. Orange rectangles represent modelling steps. Green rectangles are the results of the analysis. ITN=insecticide-treated net. IRS = Indoor Residual Spraying, SMC = Seasonal Malaria Chemoprevention. *Pf*PR=*Plasmodium falciparum* parasite rate. S1 BAU = business-as-usual scenario. S2 noPMI = scenario with PMI-funded interventions removed.

### Determining malaria control coverage attributable to PMI in 2025

The Roll Back Malaria Initiative (RBM) hosts programmatic and financial gap analysis tables that include data – by malaria-endemic country – detailing the number of ITNs financed for planned distribution in 2024 and 2025, reflecting a business-as-usual (BAU) scenario^14^. These numbers are inclusive of all sources of funding, including PMI. At the time of analysis, this information was current as of 17 December 2024. Separately, we sourced historical ITN distribution data from national malaria control programs (NMCPs) for the period 2000-2023, primarily via World Health Organization reporting^6^. We then refined the financed 2025 annual distribution volumes into inferred quarterly volumes and, where relevant, subnational target regions based on information on the planned timings and locations of mass campaigns available via the ‘ITN Mass Campaign Tracker’ resource maintained by the Alliance for Malaria Prevention^15^. The fraction of ITNs likely to be distributed through mass-campaign distributions versus routine distribution channels (for example via health facilities, antenatal clinics, schools, or community health workers) was inferred based on this ratio for each country in historical data. We then estimated for each country or subnational unit the fraction of all ITNs financed for 2025 that are financed by PMI. Data on planned volumes of ITNs procured or distributed by PMI versus other funders in 2025 were extracted from PMI Fiscal Year (FY) 2024 Malaria Operational Plans (MOPs)^16^, together with details on (i) the geographical extent of PMI support where this was subnational, and (ii) where PMI provides financial support to facilitate distribution of ITNs purchased by other donors (including in Cameroon and Nigeria). The resulting inferred PMI-financed proportions were then subtracted from the BAU volumes to define a ‘no PMI’ scenario (see appendix table S1). Next, both the BAU and no-PMI scenario ITN volumes were entered into an established dynamic coverage model^10,11^ which transformed them into monthly estimates of ITN coverage (household access and use) at subnational level for 2025. The model took into account ITNs already distributed to households in preceding years and their likely rate of loss. In the no-PMI scenario, we did not explore possible geographical reallocation of ITNs funded by other means.

Data on the volumes of ACTs financed for distribution in 2025 from all funding sources, and the fraction supported by PMI, were extracted from the PMI FY2024 MOPs. These documents also provided delineation, where relevant, of the subnational regions in which PMI provide this support. The MOPs also detail past years distributions and end-of-year stock-on-hand. Combining these data elements allowed us to generate inferred subnational ACT distribution volumes for 2025 under the BAU (i.e. including planned PMI contributions) and no-PMI (excluding those contributions) scenarios (see appendix figure s3). Stock-on-hand at the outset of 2025 was considered available for distribution even in the no-PMI scenario, although only within existing national or subnational units. We then converted these distribution scenarios into estimated treatment coverages (i.e. the fraction of clinical malaria cases receiving effective treatment) as follows. For the BAU scenario we used pre-existing coverage estimates developed by MAP for the annual WHO World Malaria Report^12,6^ that consider rates of care seeking, access to antimalarial drugs, and the time-varying efficacy of different drug classes. We used the estimates for 2024 as a proxy for 2025, assuming coverage would be similar under BAU. Treatment coverage under the no-PMI scenario in 2025 was then computed by reducing public-sector treatment rates in proportion to the fraction of all ACTs financed in 2025 by PMI per subnational unit. We did not extend this analysis to include the possible effects of reduced availability of rapid diagnostic tests (RDTs) under the no-PMI scenario. In reality, reduced access to confirmatory diagnosis would further compromise levels of access to effective antimalarial treatment by reducing appropriate targeting of limited ACT stock to confirmed cases. In the no-PMI scenario, we did not explore possible geographical reallocation of ACTs funded by other means.

Areas with planned SMC campaigns supported by PMI under BAU in 2025 were identified using data from the FY2024 PMI MOPs, augmented with data reported by NMCPs to WHO^6^. Within those areas, coverage under BAU was assumed to be the same as that estimated for 2024 based on a compilation of post-campaign coverage surveys undertaken by NMCPs and by Malaria Consortium^17,18^. The timing and number of cycles were assumed to be the same as in 2024 and the target population was assumed to be strictly children under 59 months. The no-PMI scenario was then simply defined as having zero SMC coverage in 2025 in the PMI-supported SMC areas.

The most recent MAP estimates of IRS coverage (for the year 2023) were obtained, which are based on subnational data on spray campaigns reported by NMCPs to WHO^6^, reported by PMI in national ‘End of Spray Reports’ (e.g. PMI VectorLink reports^19,20^), or from other country-specific sources^21^. We used this 2023 coverage to represent our BAU scenario for 2025. Details of the subset of planned IRS spraying supported by PMI in 2025 were extracted from PMI FY2024 MOPs^16^. The no-PMI scenario was then defined as having zero IRS coverage in 2025 in those IRS target areas supported by PMI.

For all interventions, the no-PMI scenario was defined in the absence of any mitigation strategies, such that coverage supported by PMI under BAU was assumed to be lost and not replenished by other means.

### Determining epidemiological impact of PMI-attributable coverage

We propagated the BAU and no-PMI intervention coverage scenarios for ITNs, ACTs, SMC, and IRS through a series of established models to generate corresponding estimates of infection prevalence, clinical case incidence, and malaria mortality at 5×5 km resolution and by month across 2025.

First, we fitted a Bayesian geostatistical model to historical data on malaria infection prevalence, intervention coverage, and environmental and socioeconomic covariates from 2000-2023. Prevalence data comprised 59,516 cross-sectional observations of malaria infection prevalence age-standardised to the 2-10 year old age group (i.e. *P. falciparum* parasite rate, *Pf*PR_2-10_), as compiled on a rolling basis by MAP^12^ and collected predominantly through cross-sectional surveys such as those conducted by the Demographic and Health Surveys Program (DHS)^22^. Observations were standardised for diagnostic type^23^ and age^24^ before modelling. Geotemporal intervention coverage estimates from 2000-2023 were derived from the exiting MAP coverage models referenced above and described in detail previously^13^. Geotemporal data on environmental and socio-economic covariates capturing climatic, landcover, and infrastructural variables were also derived from existing MAP catalogues as described previously^13^. The geostatistical modelling framework used here is described extensively elsewhere^5,13^. In brief, the effect on *Pf*PR_2-10_ of: (i) the physical and insecticidal components of ITN coverage, as a function of baseline transmission intensity and trends in pyrethroid resistance; (ii) access to effective antimalarial treatment; and (iii) IRS coverage was determined empirically using a hierarchical Bayesian spatio-temporal model. The model included additional terms controlling for (iv) changing environmental and socioeconomic predictors; (v) underlying baseline endemicity; and (vi) unexplained variation accounted for using a spatio-temporal random field with separable Matérn-5/2-AR(1) covariance structure and independent country-level random effects. For this analysis, we updated the model to explicitly include the effect of insecticide resistance (see appendix section 2.2) and leverage new remote-sensing imagery to better resolve urban-rural gradients of risk. Fitted parameters are shared in the appendix, tables S3 and S4. After five-fold cross-validation, out-of-sample mean squared error was 2·1%, mean absolute error was 9·0%, and correlation between observed and predicted *Pf*PR_2-10_ was 0.80. The model was fit in R v.4.4.1 using INLA v.24.06.27^25,26^. The fitted model, which includes detailed terms characterising the effects of each intervention class, was then used in a final step to predict *Pf*PR_2-10_ at 5×5 km resolution for each month of 2025 across Africa. Two predictions were carried out to reflect intervention coverages under both the BAU and no-PMI scenarios described above. Malaria’s transmission dynamics are uncertain and heterogeneous, but suggest the impact of changing intervention coverage on *Pf*PR_2-10_ would not be immediate^27^, so coverages were lagged by both one and two months, before pooling to calculate summary statistics. For both 2025 scenarios, environmental and socioeconomic covariates were fixed at 2023 levels, the most recent full year of available data.

The BAU and no-PMI *Pf*PR_2-10_ predictions for 2025 were used to generate corresponding geotemporal estimates of annual *P. falciparum* clinical incidence rate using an established natural history model^28^ that estimates age-structured clinical incidence as a function of *Pf*PR, seasonality, and exposure history. Annual estimates were disaggregated to monthly values using a seasonality model based on compiled observations of clinical malaria seasonality across Africa^29^. We additionally accounted for the impact on clinical incidence of SMC using our modelled SMC coverage combined with observations of SMC effectiveness under routine implementation pooled across sites in seven West African countries^30^ (see appendix section 2.3.1). Malaria mortality in 2025 under the BAU and no-PMI scenarios was determined by first combining the geotemporal estimates of clinical cases and (unlagged) treatment rates to derive untreated cases. We then applied to those untreated case estimates an untreated case fatality rate derived from the World Malaria Report^31^, including upper and lower bound estimates. Note that this approach captures two mechanisms by which antimalarial treatment averts mortality: via direct impact on *Pf*PR_2-10_, which reduces clinical incidence rates, and then additionally by reducing the fraction of clinical cases that progress to death.

### Role of the funding source

The funder of the study had no role in study design, data collection, data analysis, data interpretation, or writing of the report. The corresponding author had full access to all the data in the study and had final responsibility for the decision to submit for publication.

## Results

### Intervention coverage

Our compiled data on planned procurement and distribution indicated that under business-as-usual PMI would provide 44 million ITNs in 2025. An additional 15 million ITNs funded by the Against Malaria Foundation for Nigeria and Cameroon are reliant on PMI logistical support for distribution^32,33^. In total, these volumes represent 35% of the 169M ITNs planned for distribution in 2025 across the 25 African PMI focus countries included in this study. We estimated that these ITNs would contribute 10.1 percentage points (pp) to population coverage (i.e. use – the percentage of people sleeping under an ITN) by the end of 2025 across the PMI focus countries. Impact varies considerably by country with effects exceeding 20 pp in Sierra Leone, Nigeria (PMI focus states), Burundi, Zimbabwe, and Cameroon (Figure 2A). Data on planned ACT procurement and distribution indicated that PMI would finance 71 million doses of the drug across its African focus regions in 2025 under business-as-usual, representing 52% of total ACT procurement in those regions. We estimated that these drugs would contribute 12·0 pp to the fraction of clinical malaria cases receiving effective treatment. Some countries are proportionally more reliant on PMI-funded ACTs than others, with PMI contributing over 20 pp to effective treatment rates in Zambia, Senegal, Burundi, and PMI focus provinces of Angola (Figure 2B). Compiled data indicates that under business-as-usual PMI would contribute 12·3 pp to under-five population coverage of SMC across all areas targeted for this intervention in Africa in 2025. In specific subnational geographies where PMI would concentrate SMC support, this figure is often greater than 75 pp (Figure 2C). PMI was scheduled to be a relatively minor contributor to IRS campaigns in Africa in 2025, with a business-as-usual contribution of 4·9 pp to annualised coverage across regions targeted continent-wide (Figure 2D).

**Figure 2.**
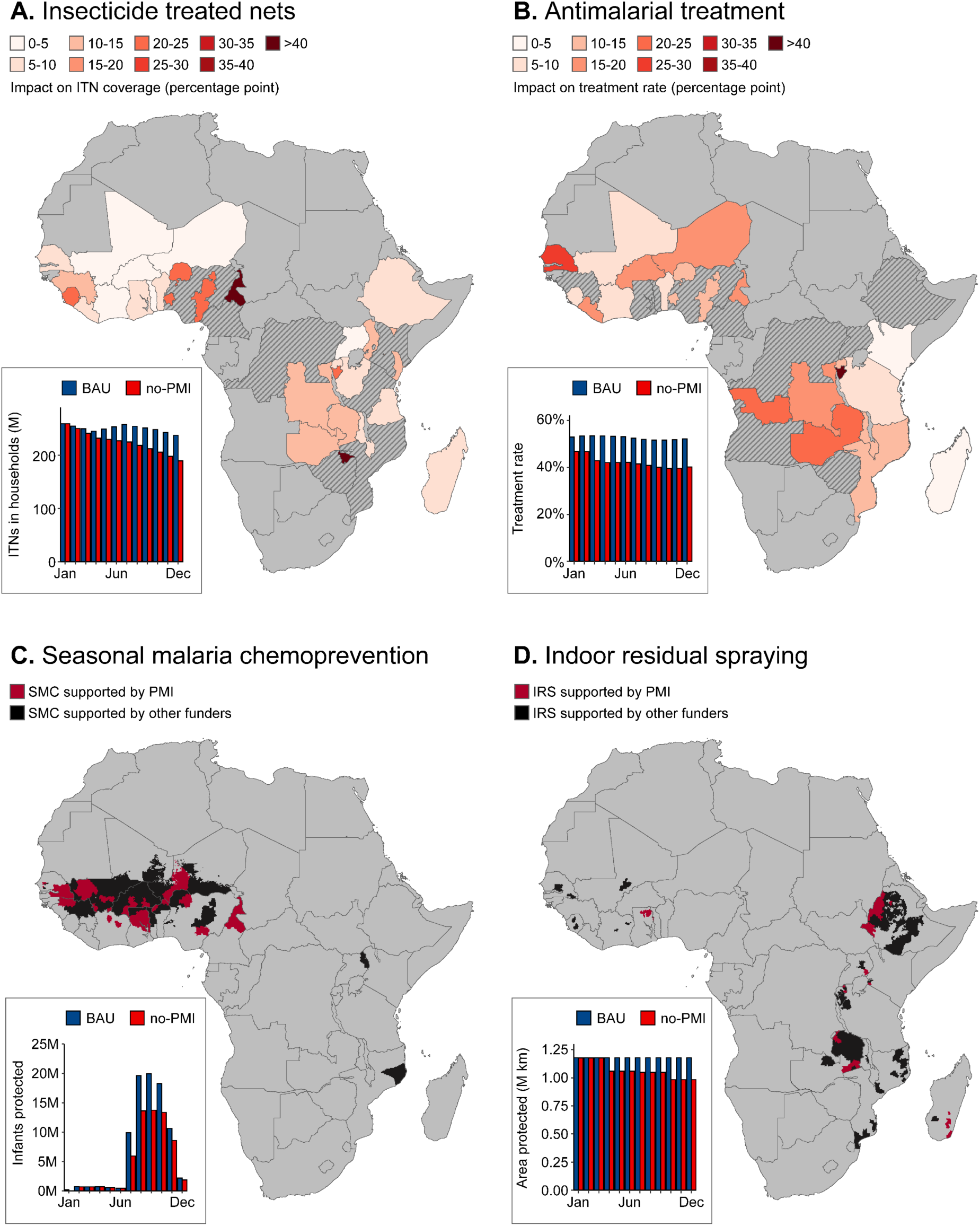
Projected contributions of PMI in 2025 under business-as-usual (BAU) to each intervention class considered in this study. (A) Percentage-point difference in access to insecticide treated bednets between BAU and no-PMI scenarios, projected in December 2025. Hashed areas correspond to regions of PMI focus countries where PMI does not procure ITNs nor support their distribution. Inset shows BAU and no-PMI scenario projected ITNs in households, January-December 2025. (B) Percentage-point difference in the proportion of clinical malaria cases receiving effective treatment between BAU and no-PMI scenarios, projected in December 2025. Hashed areas correspond to regions of PMI focus countries where PMI does not procure ACTs. Inset shows BAU and no-PMI scenario projected effective treatment rates of clinical malaria cases, January-December 2025. (C) Spatial extent of Seasonal Malaria Chemoprevention projected in 2025, classified by funding support (PMI vs other funders). Inset shows BAU and no-PMI scenario projected numbers of infants (under five) protected by SMC, January-December 2025. (D) Spatial extent of IRS projected in 2025, classified by funding support (PMI vs other funders). Inset shows BAU and no-PMI scenario projected area (in km^2^) protected by IRS, January-December 2025. SMC = Seasonal Malaria Chemoprevention; BAU = business-as-usual; PMI = President’s Malaria Initiative.

### Malaria cases and deaths

We estimate that PMI procurement and distribution of malaria control commodities, as planned under business-as-usual operations for 2025, would avert 13·6M (95% uncertainty interval 11·4M – 16·4M) clinical malaria cases and save 104,000 (69,000 – 161,000) lives across that year. (Table 1, Figure 3A,B). Burden averted by month is slightly greater in the latter half of the year, driven in part by the availability of pre-existing stock-on-hand for ACTs which reduce the impact of the no-PMI scenario in the first few months of 2025. In addition, the timing of planned PMI-funded ITN and SMC campaigns, which typically occur in quarters three and four for Sahelian countries due to seasonal variations in transmission intensity, means greater impact in these later months. Our decomposition by intervention type (Figure 3B,C) indicated that PMI-funded ACT provision formed the largest contribution to both cases (55%, 95% uncertainty interval 48 – 67%) and deaths (84% [82 – 85%]) averted, followed by ITNs (25% of cases [20 – 31%], 7% of deaths [6 – 8%]) and SMC (20% of cases [11 – 23%], 9% of deaths [8 –10%]). At national-level, we estimate business-as-usual PMI activities would save the largest number of lives in PMI focus regions of Nigeria (20,000; 14,000 - 30,000), Burundi (11,000; 7,100 - 18,000) and Democratic Republic of the Congo (11,000; 6,200 - 20,000), corresponding to densely-populated high-burden countries with extensive PMI support (Table 1).

**Figure 3.**
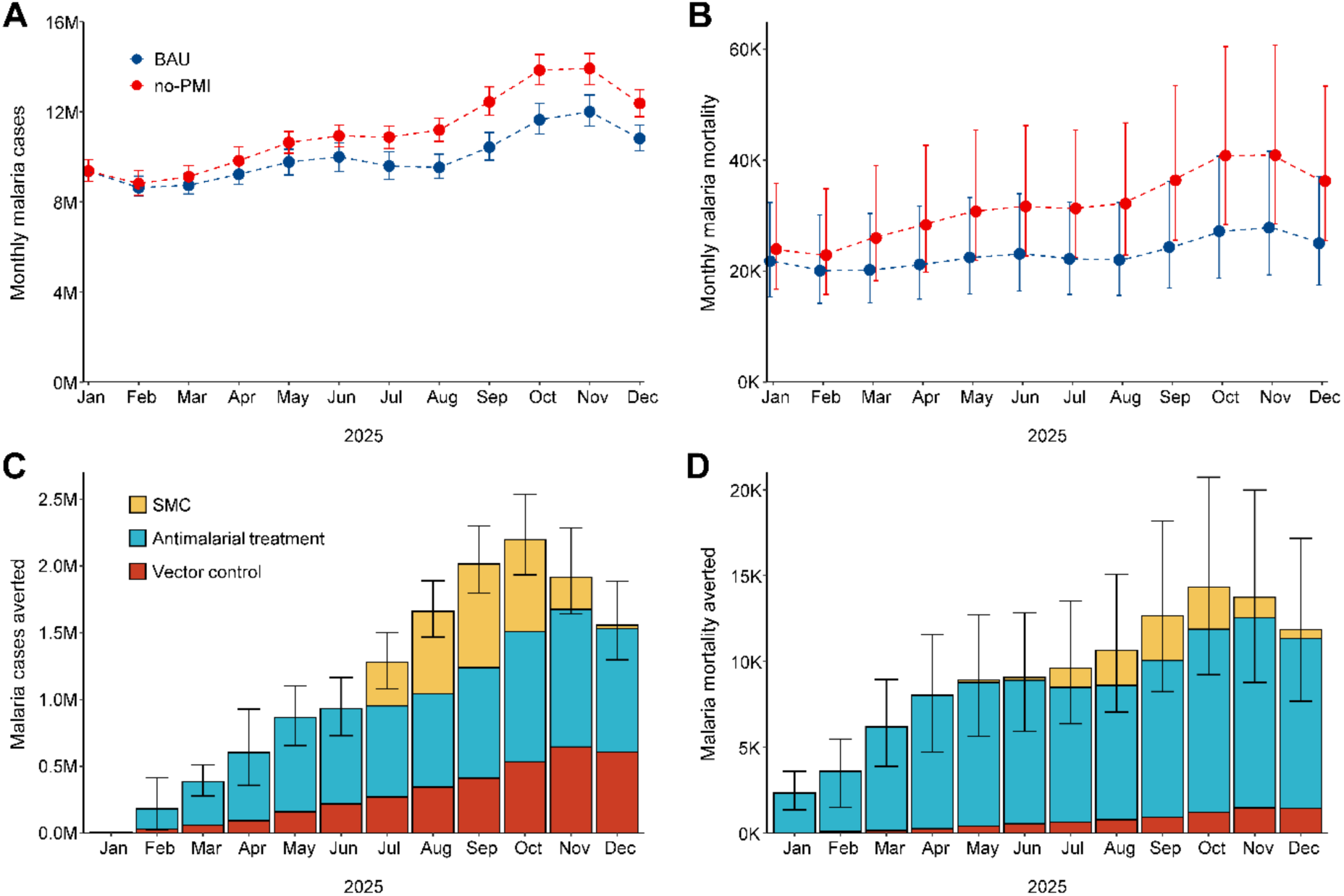
Estimates of malaria burden averted by PMI in Africa in 2025 with 95% uncertainty intervals; (A) Number of *Plasmodium falciparum* clinical cases (in millions), by month, with blue representing business-as-usual projection and red with PMI-funded interventions removed; (B) the number of *P. falciparum* deaths, by month, with colour scheme as in (A). Plotted monthly estimates are off-set in the no-PMI scenario for presentation and ease of interpretation; (C) Number of *P. falciparum* clinical cases (in millions) averted by PMI-funded interventions, by month of 2025, disaggregated by intervention class. (D) Number of *P. falciparum* deaths averted by PMI-funded interventions, by month of 2025, disaggregated by intervention class. Vector control consists of both coverage with Insecticide Treated Bednets and Indoor Residual Spraying; SMC = Seasonal Malaria Chemoprevention.

**Table 1.** Estimated mean malaria cases and deaths averted in Africa by PMI in 2025, with 95% uncertainty intervals. National level malaria case estimates are rounded to three significant figures, deaths estimates to two. Africa-wide estimates are rounded to the nearly 100,000 (cases) or 1,000 (deaths). DRC = Democratic Republic of the Congo. Africa estimate corresponds to the sum across all PMI focus geographies.

| Country | Malaria cases averted | Malaria deaths averted |
| --- | --- | --- |
| Angola | 165,000 (117,000 - 221,000) | 1,800 (1,000 - 3,200) |
| Burundi | 1,040,000 (766,000 - 1,360,000) | 11,000 (7,100 - 18,000) |
| Benin | 251,000 (217,000 - 287,000) | 1,200 (840 - 1,800) |
| Burkina Faso | 888,000 (754,000 - 1,040,000) | 7,200 (4,800 - 11,000) |
| Cote d'Ivoire | 459,000 (344,000 - 599,000) | 2,600 (1,800 - 3,900) |
| Cameroon (PMI focus) | 1,510,000 (1,350,000 - 1,650,000) | 4,400 (3,300 - 5,900) |
| DRC (PMI focus) | 1,260,000 (983,000 - 1,610,000) | 11,000 (6,200 - 20,000) |
| Ethiopia | 190,000 (150,000 - 233,000) | 400 (120 - 930) |
| Ghana | 248,000 (219,000 - 273,000) | 590 (500 - 700) |
| Guinea | 200,000 (173,000 - 225,000) | 450 (340 - 600) |
| Kenya | 148,000 (120,000 - 177,000) | 1,100 (950 - 1,400) |
| Liberia | 120,000 (88,700 - 158,000) | 1,200 (740 - 1,900) |
| Madagascar | 36,000 (26,400 - 47,900) | 220 (69 - 470) |
| Mali | 515,000 (430,000 - 618,000) | 2,800 (2,000 - 3,900) |
| Mozambique | 419,000 (276,000 - 594,000) | 4,900 (2,700 - 9,300) |
| Malawi | 312,000 (221,000 - 428,000) | 2,500 (1,800 - 3,600) |
| Niger | 770,000 (563,000 - 1,060,000) | 7,900 (5,200 - 12,000) |
| Nigeria (PMI focus) | 2,850,000 (2,490,000 - 3,280,000) | 20,000 (14,000 - 30,000) |
| Rwanda | 98,100 (71,500 - 127,000) | 690 (510 - 920) |
| Senegal | 946,000 (676,000 - 1,250,000) | 7,700 (2,700 - 17,000) |
| Sierra Leone | 171,000 (140,000 - 201,000) | 1,200 (700 - 1,900) |
| Togo | 5,350 (4,140 - 6,460) | 9 (6 - 13) |
| Tanzania | 582,000 (417,000 - 767,000) | 9,300 (7,400 - 12,000) |
| Uganda | 107,000 (91,100 - 125,000) | 140 (100 - 230) |
| Zambia | 298,000 (218,000 - 402,000) | 3,300 (2,500 - 4,600) |
| Zimbabwe | 6,940 (4,820 - 9,310) | 20 (6 - 47) |
| **Africa (PMI focus)** | **13.6M (11.4M – 16.4M)** | **104,000 (69,000 – 161,000)** |

## Discussion

At a time of ongoing policy reorientation, it is essential to make available to decision makers reliable measures of the impact of investments in global health. Here we demonstrate that a fully functioning PMI would likely avert nearly 14M malaria cases and around 100,000 malaria deaths in Africa in 2025. These estimates do not rely on simple extrapolations of lives-saved- per-dollar but are instead built up from a detailed reconstruction of planned procurement and distribution activities for the major classes of malaria control commodity, the proportional geographically-specific contribution of PMI to that commodity provision, the way those commodities drive population coverage, and the way that coverage impacts malaria transmission and burden. This granularity allows the necessary sophistication to capture the differential effects of each intervention class, to take into account control interventions already in place or available for distribution at the onset of the year, and to acknowledge heterogeneity in underlying malaria risk across the continent and how this influences intervention impact.

The geospatial methodology used in this analysis does not explicitly account for immune rebound and associated hysteresis in intervention impact. Rebound to above pre-intervention burden levels of burden – due to the suppression of naturally acquired immunity – has been observed in settings where sustained high-frequency chemoprophylaxis has been abruptly removed^34^, but the frequency and magnitude of this effect relative to resurgence to pre-intervention levels is poorly understood^35,36,37^. A review of fifty studies suggested immune rebound occurs infrequently (8/50 studies) under trial conditions^35^. Under real-world conditions intervention coverage fluctuates (appendix Figures S5, S6), predominantly due to cyclic vector control campaigns^11^, leading to periods of reduced protection and increased exposure. The extent to which the no-PMI scenario – a partial reduction of imperfect coverage –could lead to a rebound effect is therefore unclear, but in this respect our morbidity and mortality estimates may be conservative.

The results presented here must be interpreted in the context of several limitations. First, while we focus on PMI-funded commodity procurement, we do not explicitly account for PMI support to logistics and supply chain management, instead assuming that changes in coverage stem only from reduced input commodity volumes. Second, we do not account for PMI support for RDTs. In practice, RDT stock-outs would result in reversion to presumptive malaria diagnosis which would translate into a higher fraction of ACTs being prescribed to non-malaria cases. This would not only impair appropriate management of non-malaria causes but would place further strain on ACT stocks. Reduced confirmatory diagnosis also means surveillance data would decline in quality – impairing future monitoring of malaria trends. Third, when estimating mortality effects, we take into account only changes to overall case incidence and to rates of first-line treatment of presenting uncomplicated clinical cases, implicitly assuming the case-fatality rate among untreated cases remains constant. This neglects any contribution of PMI to supporting quality management of severe malaria cases - for example via procurement of injectable artesunate. Fourth, we do not account for any present or planned PMI support for the implementation of vaccination with the RTS,S/AS01 or R21/Matrix-M vaccines. Fifth, we do not account for healthcare personnel directly supported by PMI. Most importantly, this includes around 100,000 community health workers^7^ whose efforts have demonstrably improved access to timely malaria case management in rural and remote communities in many PMI focus countries^38^. Sixth, we do not account for a range of additional activities supported by PMI that include strengthening of disease and entomological surveillance systems, and monitoring of insecticide and drug resistance. Seventh, we do not capture the indirect effects of increased malaria burden on health systems, and its potential to reduce capacity for care of other conditions.

It is important to note that nearly all the limitations listed above imply our estimates are conservative with respect to the number of cases and deaths averted. Conversely, we do not consider possible mitigation strategies in a scenario without PMI support – instead focusing on PMI impact *ceteris paribus*. This likely neglects the resilience and increasing self-sufficiency of African health agencies in many countries. In reality, a future with more constrained international funding for malaria control would catalyse a range of mitigation and response strategies by national government and health agencies. These could include emergency domestic funding mechanisms, amplifying the existing trend towards greater domestic funding for malaria control^6^. Limited control commodities could also be reprogrammed - either geographically or across different intervention mixes – to maximise disease control within a given resource envelope. However, the degree of readiness and the time required for these structural responses to be enacted is likely to vary considerably across PMI focus countries. Furthermore, given that PMI contributes nearly a quarter of the total $3.0 billion USD of malaria funding spent in the WHO AFRO region^6,7^, rapidly mitigating the absence of PMI at the necessary scale would require significant reallocation of resources from other budgets. A default scenario without substantial mediation remains, therefore, an appropriate benchmark to consider for the forthcoming year. Compared to this scenario, our analysis indicates that PMI investment in supporting procurement and distribution of malaria control commodities would translate directly into millions of malaria cases averted and a hundred thousand lives saved in its 27 countries of operation in Africa across 2025.

## Contributors

PWG and TLS conceptualised the study, led data analysis and wrote the first draft of the manuscript. JL, MM, AS, MV, HB, YB, SH, RJ, PA, AB, EC, CV, SFR, NG, DJW contributed to data curation and compilation and contributed to data analysis. All authors had full access to the data, contributed to review and approved the final version of the manuscript. The corresponding author had full access to all the data in the study and had final responsibility for the decision to submit for publication.

## Data sharing

Data on commodity funding and distribution, malaria prevalence and intervention coverage used in this analysis are available in the public domain via the cited sources. All PMI-focus-geography level results are available within the main text and supplementary material. Other summarisations are available upon request to the authors.

## Declaration of interests

All authors declare no competing interests.

## Acknowledgments

This work is supported by the National Health and Medical Research Council, Australia (2025280).

## Notes

### Competing Interest Statement

The authors have declared no competing interest.

### Summary of Updates

Estimates have been updated. Results and figures have been revised.

